# Is Nigeria really on top of COVID-19? Message from effective reproduction number

**DOI:** 10.1101/2020.05.16.20104471

**Authors:** A. I. Adekunle, O. A. Adegboye, E. Gayawan, E. S. McBryde

**Affiliations:** Australia Institute of Tropical Health and Medicine, James Cook University, Townsville; Decision and Modelling Science, Victoria University, Melbourne; Biostatistics and Spatial Statistics Research Group, Department of Statistics, Federal University of Technology, Akure

**Author notes:** Corresponding Author’s.

**Keywords:** Covid-19, Nigeria, effective reproductive number, basic reproduction number, imported cases

## Abstract

Following the importation of Covid-19 into Nigeria on the 27 February 2020 and then the outbreak, the question is: *how do we anticipate the progression of the ongoing epidemics following all the intervention measures put in place?* This kind of question is appropriate for public health responses and it will depend on the early estimates of the key epidemiological parameters of the virus in a defined population.

In this study, we combined a likelihood-based method using a Bayesian framework and compartmental model of the epidemic of Covid-19 in Nigeria to estimate the effective reproduction number (*R*(*t*)) and basic reproduction number (*R*_0_). This also enables us to estimate the daily transmission rate (*β*) that determines the effect of social distancing. We further estimate the reported fraction of symptomatic cases. The models are applied to the NCDC data on Covid-19 symptomatic and death cases from 27 February 2020 and 7 May 2020,.

In this period, the effective reproduction number is estimated with a minimum value of 0.18 and a maximum value of 1.78. Most importantly, the *R*(*t*) is strictly greater than one from April 13 till 7 May 2020. The *R*_0_ is estimated to be 2.42 with credible interval: (2.37 – 2.47). Comparing this with the *R*(*t*) shows that control measures are working but not effective enough to keep R(t) below one. Also, the estimated fractional reported symptomatic cases are between 10 to 50%.

Our analysis has shown evidence that the existing control measures are not enough to end the epidemic and more stringent measures are needed.

## Introduction

The emerging severe acute respiratory syndrome coronavirus 2 (SAR-CoV2) causing Covid-19 is still causing havoc globally. The outbreak was declared a pandemic by the World Health Organisation (WHO) on 11 March 2020. As of 2 May 2020, over 3.5 million people were confirmed to be infected with this respiratory disease, and about a quarter of a million deaths have been reported globally (1). Many countries have put intervention measures in place to contain the virus and progress has been reported in many countries (2). Despite the reported successes, the pandemic is far from being over at a global scale.

Nigeria can be said to be in the early stage of Covid-19 dynamics (3). The first case of Covid-19 importation to the country was reported on 27 February 2020 and since then, there have been many more local and imported cases (4). Following this outbreak, the Nigerian government placed a ban on international flights to prevent seeding from other affected countries and introduce lockdown and social distancing to reduce local transmissions. However, the effectiveness of these measures in Nigeria is yet to be determined.

An important parameter that is crucial for understanding the dynamics of any infectious disease is the effective reproduction number (*R*(*t*)) which must be reevaluated as the pandemic progresses to determine whether or not there is progress in containing the situation (5). The *R*(*t*) is related to the basic reproduction number, which is defined as the average number of secondary cases per typical infected case in a fully susceptible population. As epidemic progresses, the susceptible pool decreases and hence, *R*(*t*) *= R*_0_*x*, where *x* is the fraction of the population that is susceptible to infection. *R*(*t*) is also influenced by control measures to mitigate or eradicate infectious diseases. These quantities *R*(*t*) *and R*_0_, can help characterize the growth rate of an epidemic, inform recommendation for control measures and determine the impact of control measures. When *R*(*t*) *<* 1, some progress has been made and the outbreak is about to end. Otherwise, the additional measure is needed as the epidemics will continue to grow.

To understand the dynamics of Covid-19 in Nigeria, we used the likelihood-based model that was previously introduced for the Ebola pandemic (6) and adopted for Covid-19 (7) to estimate *R*(*t*) in a Bayesian framework. The approach adjusts for the contribution of imported cases and report fraction of the transmission process.

## Material and Methods

### Data sources

Daily counts of confirmed Covid-19 cases were extracted from publicly available data provided by the Nigeria Centre for Disease Control (NCDC) (4). The NCDC daily situation reports include the number of local and imported cases. From 11 April 2020, all reported cases were classified as local cases. This suggests that the travel ban imposed by the Nigerian Government on 20 March 2020 has been effective. However, due to border porosity in Africa (8), there is a high likelihood that some of the reported cases after March 20 were imported cases.

### Effective reproduction number estimation

We adopt a likelihood-based approach that allows for the evaluation of the contributions of imported cases of Covid-19 (6, 7, 9). The locally acquired infections are assumed to be Poisson distributed defined as follows:

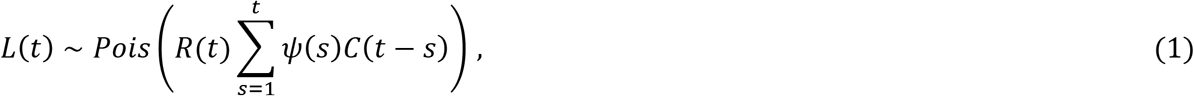

where *C*(*t*) is the number of daily new cases (locally acquired and importations) on day *t = 0, …,T. ψ*(*s*) is the distribution of the serial interval. We assumed that *ψ*(*s*) is lognormal distributed with a mean of 4.7 days and a standard deviation of 2.9 days (10). The likelihood 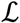 of the observed times series of cases from day 1 to day T conditional on *C*(0) is thus given by:

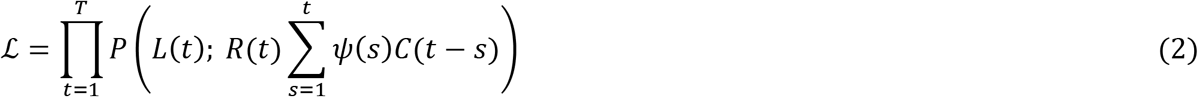

where *P* is the probability mass function of a Poisson distribution. The effective reproduction is estimated in a Bayesian framework using the Metropolis-Hasting Markov chain Monte Carlo (MCMC) method for sampling from the prior distribution of *R*(*t*). We assumed a non-informative prior, Uniform (0,10) for *R*(*t*), and performed 1,000,000 iterations with a burn-in of 10%.

An effective reproduction number aims to show how effective are the intervention strategies. The interventions put in place in Nigeria include disease surveillance to capture importations and local transmission, travel ban, contact tracing, isolation, quarantine, and social distancing (4). With a score of approximately 40% on the WHO Joint External Evaluation (JEE) mission report for Nigeria’s capacity to prevent, detect, and rapidly respond to public health risk (11), these metrics do not favour Nigeria’s Covid-19 outcomes. Therefore, to obtain the proportion of symptomatic in the daily Covid-19 cases reporting, we applied the delay-adjusted cases fatality ratio (12) to estimate the under-reporting model. Detail explanation of the model can be found in Riou, et al. (12). These metrics are adjusted as follows:

1. All importations are equally likely to cause infection as local cases.
2. All importations only contributed 40% to transmission. Here, 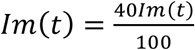. This is saying that the imported cases contributed less to the transmission and *Im*(*t*) is the number of imported cases per day.
3. Using the estimated reporting proportion, we interpreted it as reporting probability to adjust for the locally acquired cases as:

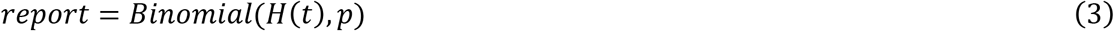 Here, *H*(*t*) is the adjusted locally acquired cases, *H*(*t*) is the cumulative reported locally acquired cases and *p* is the reporting success. For each day, we generate 1000 random variables, and for *H*(*t*) in equation (3). The median estimate is the adjusted locally acquired case. Also, previous work has shown that 50% of Covid-19 cases are asymptomatic (13). Hence, we doubled the reported cases and estimate *H*(*t*) and *R*(*t*) for 1 and 2 above.

### Modelling the dynamics of Covid-19

To forecast the number of Covid-19 cases, we estimate the basic reproduction number (*R*_0_) instead of *R*(*t*) using a compartmental model which was fitted in a Bayesian framework to the cumulative reported cases and deaths data. The compartmental model is a general SEIR-type model (S-susceptible, E – exposed, I – infectious, and R- recovered). In the model, we divide the population into ten compartments (see Table 1). The deterministic form of the model is given below:

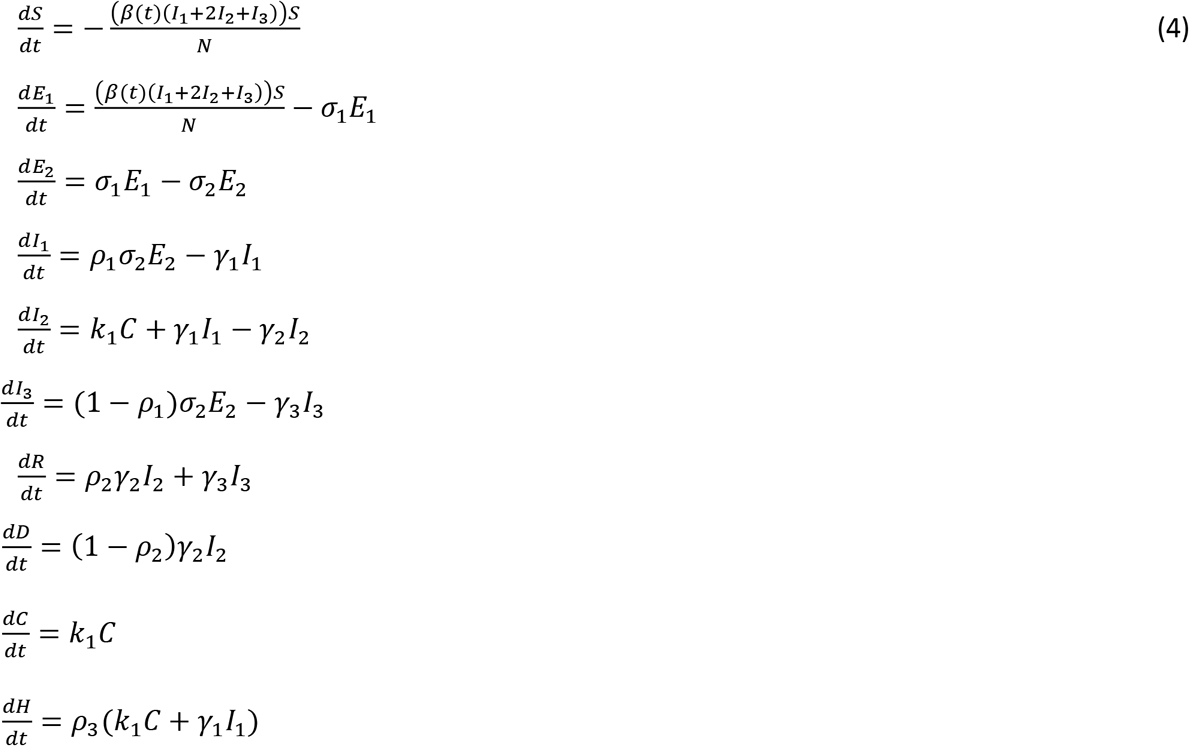

The basic reproduction number estimated using the next-generation method(14) is given as:

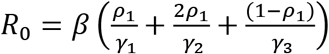

The model parameters and estimates are described in Table 2.

**Table 1:** State variable description

| Variable | Description |
| --- | --- |
| $S$ | Susceptible individuals |
| $E_1$ | Exposed individuals |
| $E_2$ | Individuals in the pre-symptomatic transmission stage during the incubation period |
| $I_1$ | Individuals that are between onset symptoms and presentation to health care stage |
| $I_2$ | Individuals with confirmed Covid-19 case and infectious |
| $I_3$ | Individuals asymptomatic |
| $R$ | Recovered class |
| $D$ | Individuals who died from Covid-19 |
| $C$ | Cumulative Covid-19 importations |
| $H$ | Cumulative symptomatic Covid-19 cases |

**Table 2:**
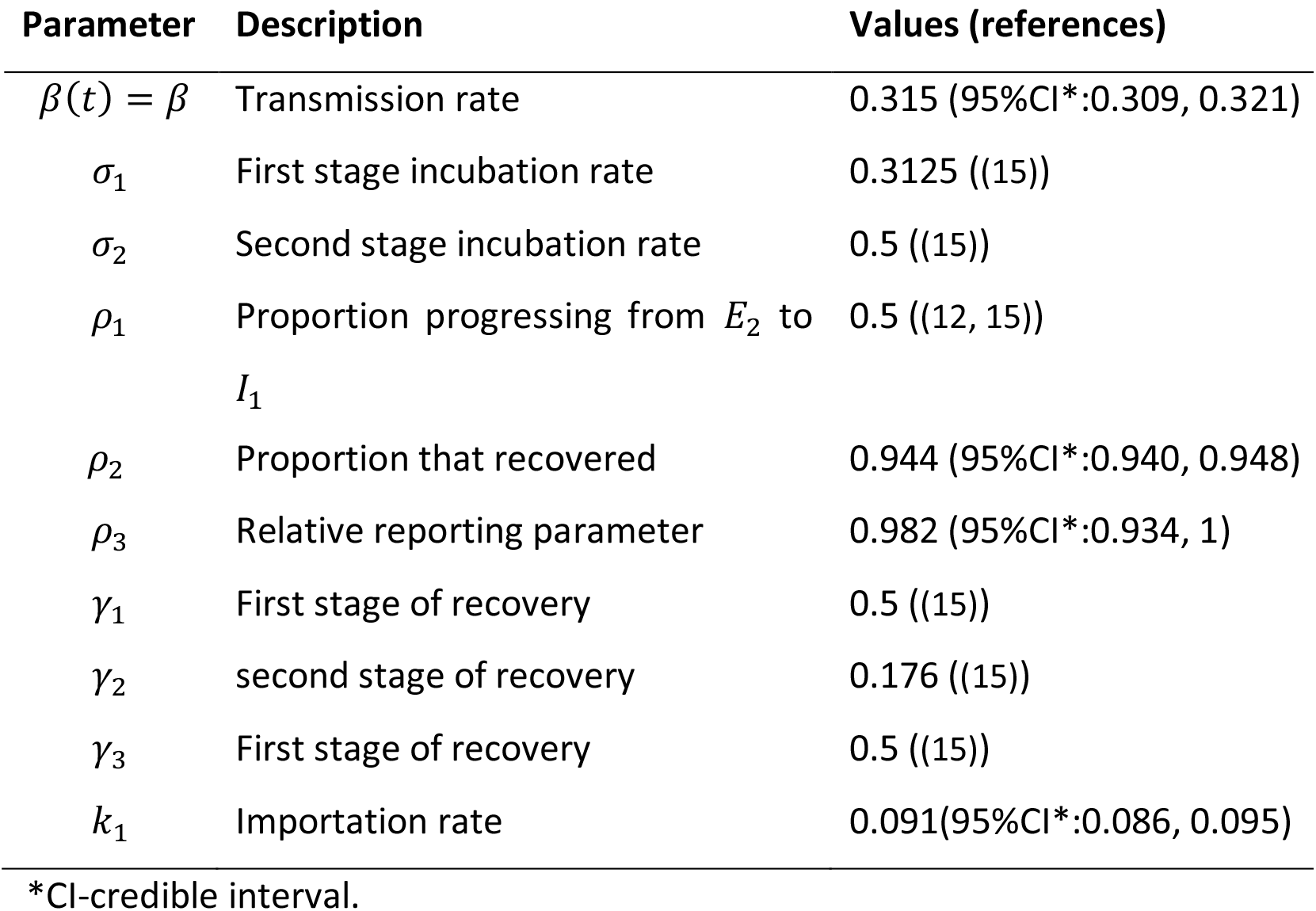
parameter descriptions

| Parameter | Description | Values (references) |
| --- | --- | --- |
| $\beta(t) = \beta$ | Transmission rate | 0.315 (95%CI*:0.309, 0.321) |
| $\sigma_1$ | First stage incubation rate | 0.3125 ((15)) |
| $\sigma_2$ | Second stage incubation rate | 0.5 ((15)) |
| $\rho_1$ | Proportion progressing from $E_2$ to $I_1$ | 0.5 ((12, 15)) |
| $\rho_2$ | Proportion that recovered | 0.944 (95%CI*:0.940, 0.948) |
| $\rho_3$ | Relative reporting parameter | 0.982 (95%CI*:0.934, 1) |
| $\gamma_1$ | First stage of recovery | 0.5 ((15)) |
| $\gamma_2$ | second stage of recovery | 0.176 ((15)) |
| $\gamma_3$ | First stage of recovery | 0.5 ((15)) |
| $k_1$ | Importation rate | 0.091(95%CI*:0.086, 0.095) |

\*CI-credible interval.

The analysis was done R (16) and the codes for the estimation can be download from https://github.com/Shina-GitHub/Covid-19_Nig.

## Results

### Epidemic data

Table 3 presents the summary statistics of the burden of Covid-19 epidemics in Nigeria from 27 February to 7 May 2020. During this period, there was a total of 3526 local cases, 210 imported cases, and 107 deaths. The death counts bring the case-fatality rate to 3.03%, a reduction in what was reported for the first 45 days of the epidemics (3).

**Table 3:** Descriptive statistics of the epidemic data of Covid-19 in Nigeria between February 27 and April 26

| Variable | Mean | SD | Minimum | Q1* | Median | Q3* | Maximum |
| --- | --- | --- | --- | --- | --- | --- | --- |
| Local cases | 73.93 | 88.53 | 1.00 | 10.00 | 32.00 | 112.50 | 381.00 |
| Imported cases | 6.10 | 4.31 | 1.00 | 2.00 | 5.00 | 9.00 | 17.00 |
| Death cases | 0.87 | 2.29 | 0 | 0 | 0 | 0 | 17 |

Q1\*- first quartile, Q3\*- third quartile

### Baseline effective reproduction number

We first estimate the effective reproduction number assuming there was no reporting delay (reporting proportion was 100%) and all the imported cases were as infectious as the locally acquired cases. The first local case (a contact of the index case) was reported on 9 March 2020, about 11 days after the importation of the first oversea case. Figure 1 (A & B) shows the baseline effective reproductive number and the model fit. The Figure reveals that the effective reproduction number was below one for the first three weeks since the introduction and increased above the threshold of one thereafter (light green box). This implies that the mitigation strategy is not optimally controlling the disease as *R*(*t*) is gradually increasing. The minimal value estimation of *R*(*t*) was 0.18 and the maximal value was 1.78. The corresponding model fit and bar graphs of the reported local and imported cases are shown in Fig 1. B

**Fig. 1.**
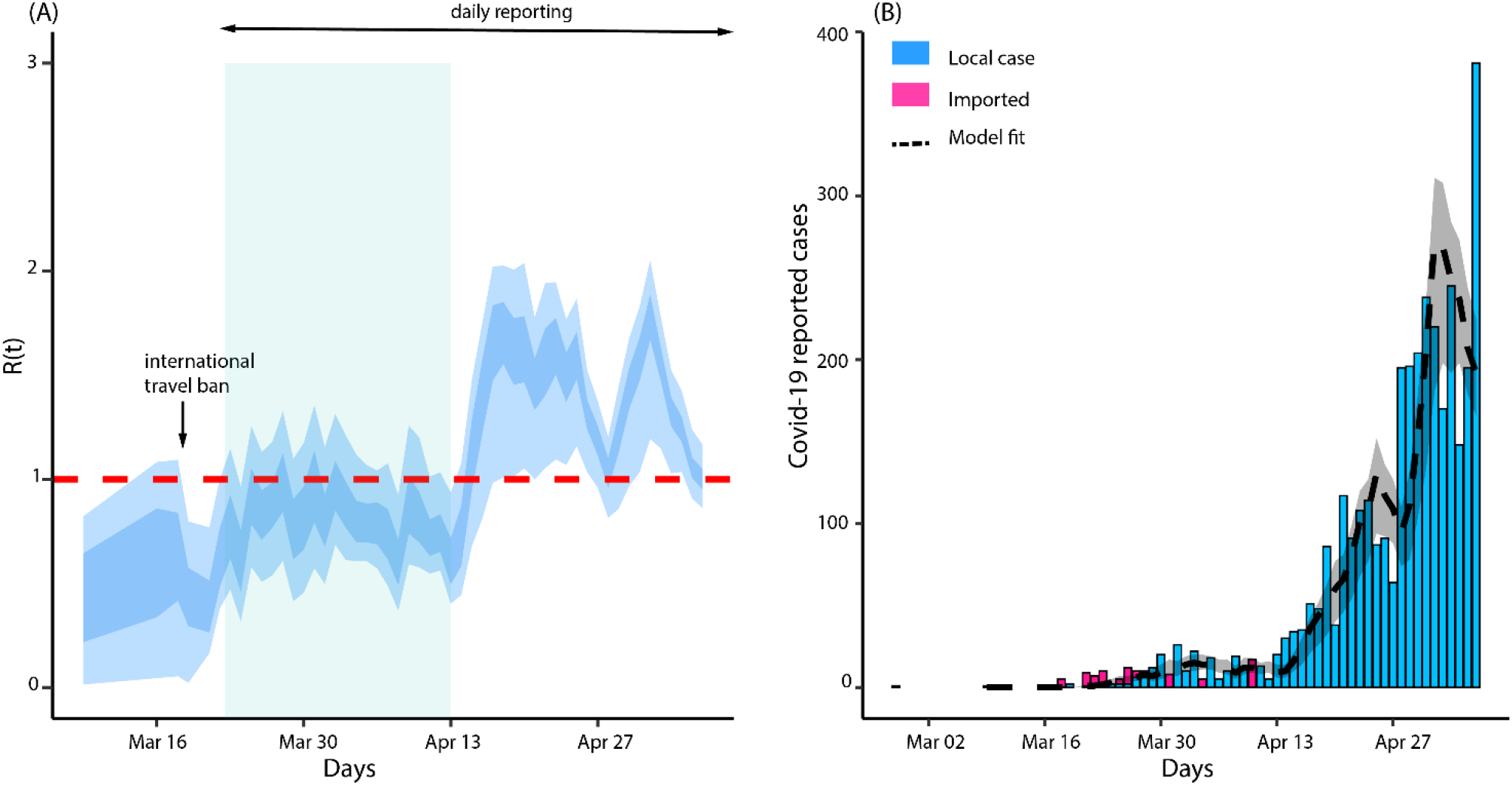
(A)Baseline effective reproduction number. The light and dark blue show 95% and 50% credible interval respectively. **(B) *R*(*t*)model fit**. The black dashed line is the median estimation of the local cases and the grey area shows a 95% credible interval of the estimated local cases.

We further estimate *R*(*t*) by assuming that 40 % of the imported cases contributed to transmission (Fig. 2A). The R(t) estimates showed the same pattern as the baseline model except that R(t) was further above one in the first three weeks since the first local case was reported (light green box (Fig. 2A)). This difference was more pronounced for the credible bands when compared to baseline estimation (see the shaded region in Fig. 1A versus Fig. 2A).

**Fig. 2.**
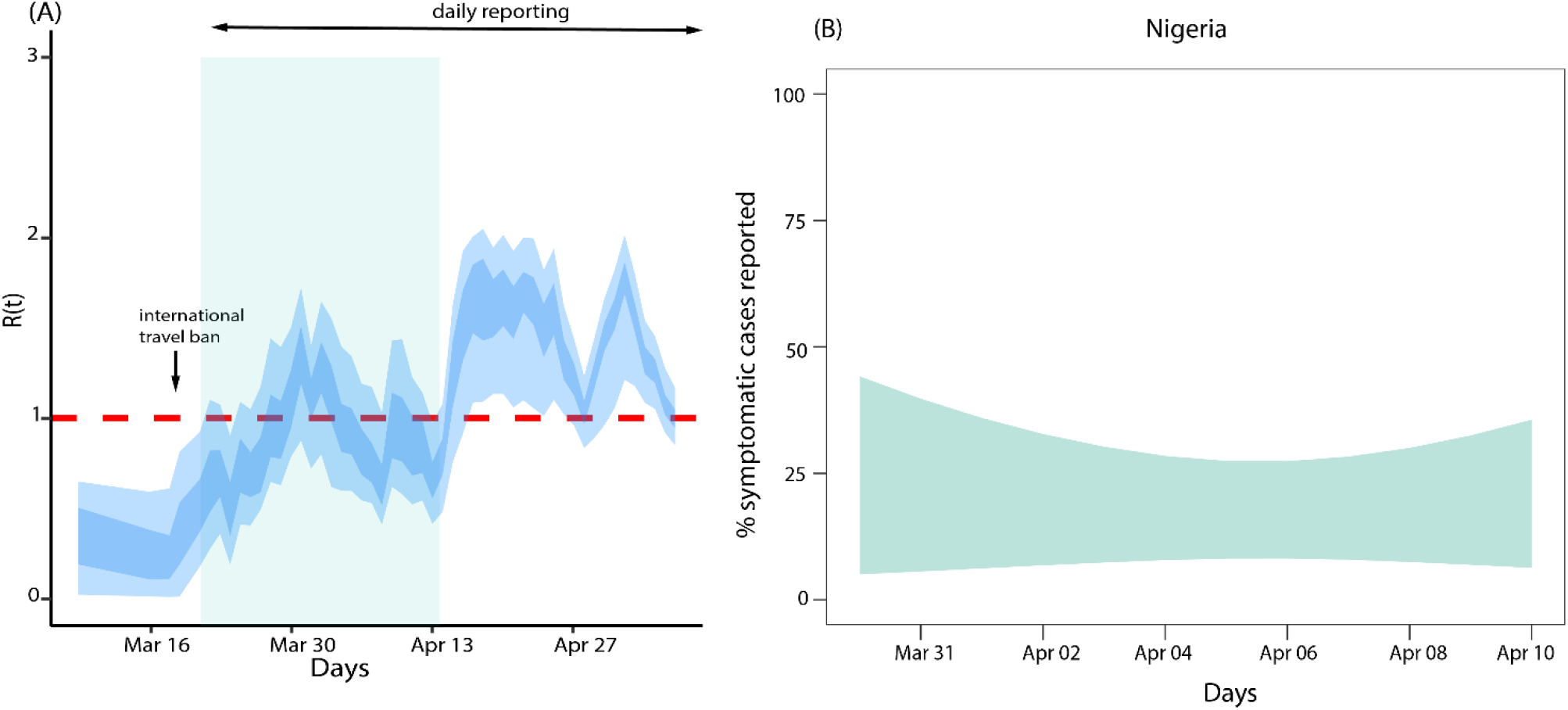
(A) Adjusted effective reproduction number for Nigeria Covid-19 epidemics based on 40% of imported cases contributing to transmission, (B) Covid-19 symptomatic reporting measure.

### Effects of symptomatic reporting proportion on effective reproduction number

We adopted the model developed by Russell, et al (13) to estimate the Covid-19 symptomatic reporting proportion for Nigeria (Fig. 2B). The results show that the under-reporting in the reported cases ranged from 10% to 50% in the two months of the outbreak. The estimate is quite large suggesting that many potential infected cases are in the community are not being reported. Again, using equation (3), we estimated *R*(*t*) to determine the effect of underreporting. First, we double the number of reported local cases to adjust for asymptomatic infection (12) and then use equation (3) to further adjust for under-reporting. The estimated *R*(*t*) was found to be above one for majority of the time considering a 10% reporting proportion (Fig. 3A) versus 50% reporting proportion (Fig 3B). These estimates were just a little higher than the baseline estimation with or without relative infectiousness of the reported cases.

**Fig. 3.**
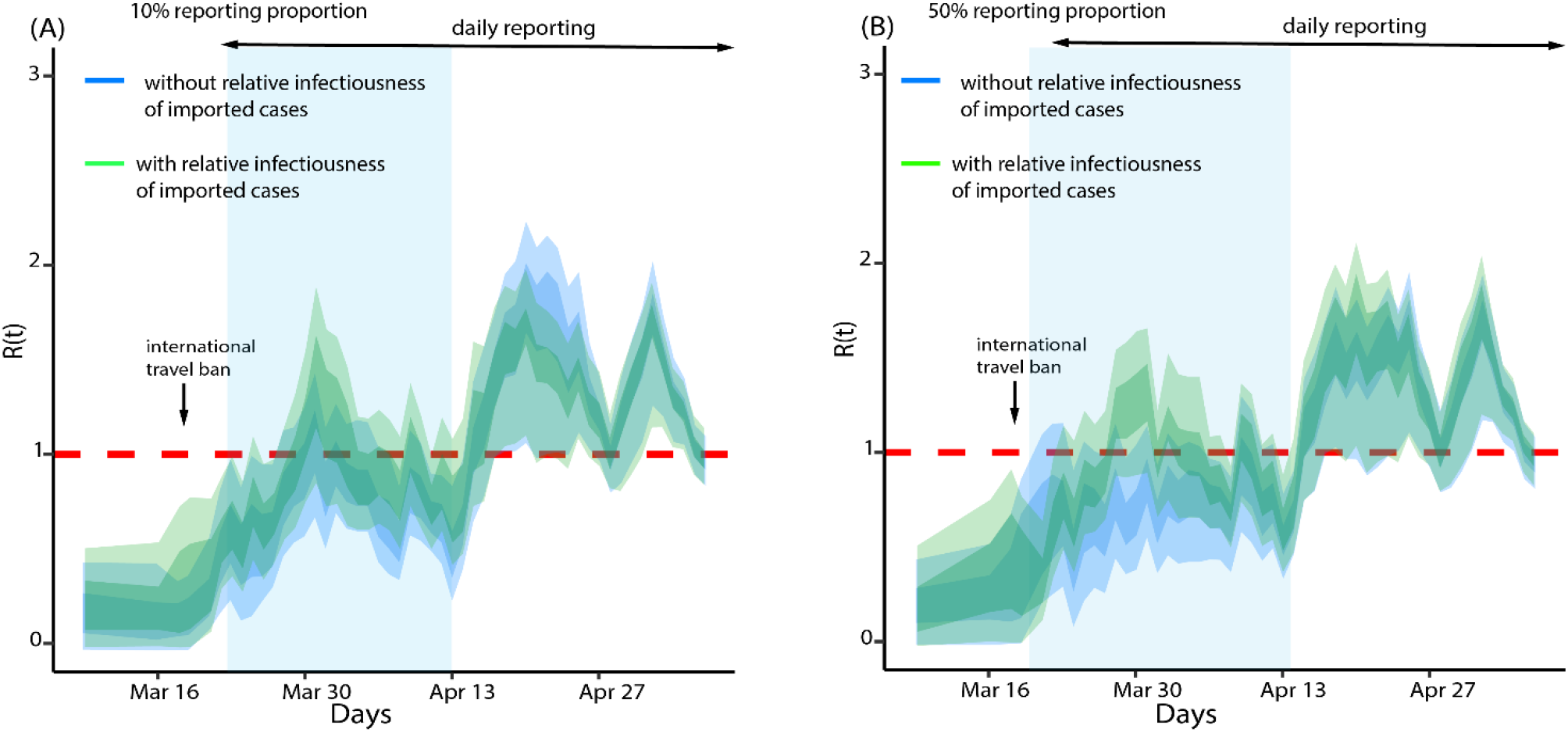
Effect of reporting proportion on effective reproduction number. (A) 10% reporting proportion (B) 50% reporting proportion.

### Model dynamics and scenario analysis

Using the compartment model (4) we estimate the basic reproduction number *R*_0_ and forecasted the cumulative number of Covid-19 cases in Nigeria for 3 weeks ahead assuming that the present dynamics is maintained. The estimated *R*_0_ was 2.42 (95% credible interval: 2.37-2.47). This estimate was similar to the *R*_0_ estimated for Wuhan during the early dynamics of Covid-19 in China (17, 18). The 3-week ahead forecasts for predicted cumulative reported number of cases and deaths are shown in Figure 4 (A and B) respectively. Our model predicted a 95% credible interval of 58890 and 69739 for cumulative reported cases by 28 May 2020 and between 1953 and 2238 for cumulative reported death cases.

**Fig. 4.**
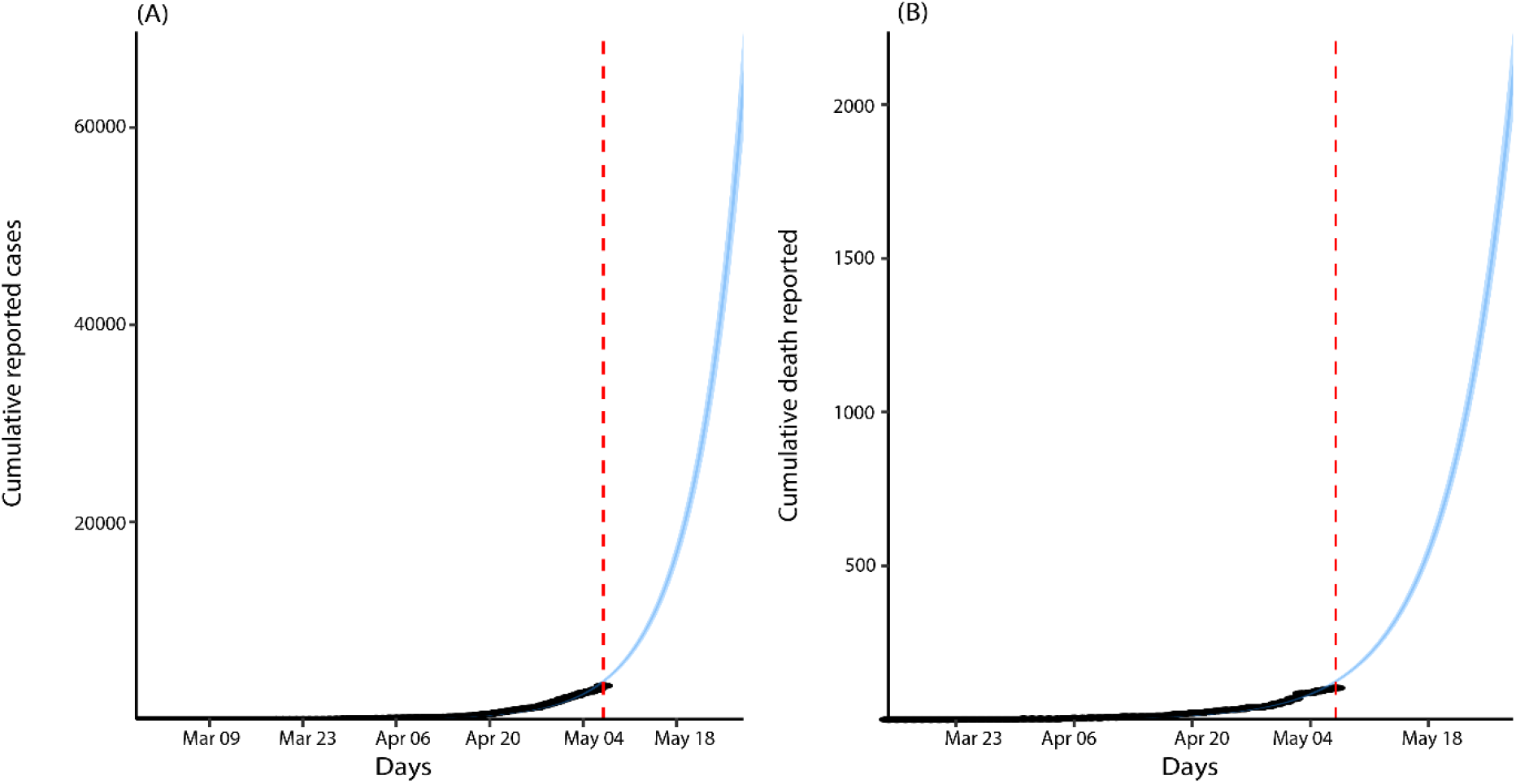
Model fit and (A) forecast of the cumulative reported cases, (B)Covid-19 deaths.

## Discussion

In this study, we estimate the effective reproduction number and basic reproduction for Covid-19 epidemic in Nigeria for the period between 27 February 2020 and 26 April 2020. Our modelling results showed that the trajectory of Covid-19 pandemic in Nigeria is far from over as the effective reproduction number is above the threshold of one and increasing. This indicates that the mitigation measures put in place have not been effective enough and efforts must be concentrated on ensuring that the existing measures are improved and additional measures that synergize the existing ones can be introduced. We estimate the basic reproduction number of Covid-19 in Nigeria is 2.63. This is similar to what was estimated for China in the early phase of the pandemic (17). With this estimate, it is clear that the mitigation measures are not able to control the disease to the level that will flatten the curve or quash it. We further determine the fraction of symptomatic proportions reported using existing work. This clearly shows that the burden of Covid-19 in the country may be more than reported.

The results presented in this study should be interpreted as a guild line with the following caveats in mind. First, imported cases are not reported daily. Considering the porosity of Nigeria borders, we expect more imported cases to be coming from other neighbouring African countries by land (8, 19, 20). Inadequate reporting may affect our estimates as we might have overestimated the burden of the pandemic because more imported cases that have not been counted probably contributed to the transmission. Under-reporting of local cases also occurs (proportion symptomatic showed that the reporting rate is between 10% and 50%.), however, this is less likely to affect our estimate, because underreporting of local cases will have a multiplier effect on both force of infection and new cases, which cancels out for the estimation of the effective reproduction number. The difference between imported and local cases explains why our estimate of effective reproduction in Figure 1A is far different from the one in Figure 3. Second, the role of asymptomatic patients in the dynamic of Covid-19 has not been fully understood (21, 22). There is evidence that these group of people contributes to the transmission process (12, 21, 22). Though we included them in our compartmental model, the likelihood model does not adjust for asymptomatic patients. This may impair the estimates as what we thought as under-reporting may be many asymptomatic patients who are fueling transmission and cannot be accounted for unless widespread testing and isolate is conducted. Lastly, the serial interval used in our estimation is derived from infector and infectee data for Japanese patients (10). This may differ in the Nigerian cohort as variability in innate immune response may affect whether a patient shows symptoms or not. Hence, getting the right pool of patients to characterize the serial interval can be challenging (23).

## Conclusion

Information derived when key epidemic parameters are estimated is important for assessing existing control strategies, adopting more effective alternatives and in policy-making. We have combined existing models with our new model to characterize the pandemic of Covid-19 in Nigeria. This work indicates that more control measures are needed to be able to stop this virus in this pandemic setting. We hope our work can guide public health responses in the fight against Covid-19 in Nigeria.

## Data Availability

Data will be provided on request

